# Does anyone fit the average? Describing the heterogeneity of pregnancy symptoms using wearables and mobile apps

**DOI:** 10.1101/2024.04.26.24306455

**Authors:** Sarah Goodday, Robin Yang, Emma Karlin, Jonell Tempero, Christiana Harry, Alexa Brooks, Tina Behrouzi, Jennifer Yu, Anna Goldenberg, Marra Francis, Daniel Karlin, Corey Centen, Sarah Smith, Stephen Friend

**Affiliations:** 4YouandMe, Seattle, WA, USA; Department of Psychiatry, University of Oxford, Oxford, UK; Computer Science Department, University of Toronto, Toronto, ON, Canada; Hospital for Sick Children, Toronto, ON, Canada; Vector Institute, Toronto, ON, Canada; Department of Laboratory Medicine and Pathobiology, University of Toronto, ON, Canada; CIFAR AI Chair, Vector Institute, Toronto, ON, Canada; Genece Health, San Diego, USA; MindMed, Inc., New York, NY, USA; Tufts University School of Medicine, Boston, MA, USA; Bodyport Inc, San Francisco, CA, USA

## Abstract

Wearables, apps and other remote smart devices can capture rich, objective physiologic, metabolic, and behavioral information that is particularly relevant to pregnancy. The objectives of this paper were to 1) characterize individual level pregnancy self-reported symptoms and objective features from wearables compared to the aggregate; 2) determine whether pregnancy self-reported symptoms and objective features can differentiate pregnancy-related conditions; and 3) describe associations between self-reported symptoms and objective features. Data are from the Better Understanding the Metamorphosis of Pregnancy study, which followed individuals from preconception to three-months postpartum. Participants (18-40 years) were provided with an Oura smart ring, a Garmin smartwatch, and a Bodyport Cardiac Scale. They also used a study smartphone app with surveys and tasks to measure symptoms. Analyses included descriptive spaghetti plots for both individual-level data and cohort averages for select weekly reported symptoms and objective measures from wearables. This data was further stratified by pregnancy-related clinical conditions such as preeclampsia and preterm birth. Mean Spearman correlations between pairs of self-reported symptoms and objective features were estimated. Self-reported symptoms and objective features during pregnancy were highly heterogeneous between individuals. While some aggregate trends were notable, including an inflection in heart rate variability approximately eight weeks prior to delivery, these average trends were highly variable at the n-of-1 level, even among healthy individuals. Pregnancy conditions were not well differentiated by objective features. With the exception of self-reported swelling and body fluid volume, self-reported symptoms and objective features were weakly correlated (mean Spearman correlations <0.1).

High heterogeneity and complexities of associations between subjective experiences and objective features across individuals pose challenges for researchers and highlights the dangers in reliance on aggregate approaches in the use of wearable data in pregnant individuals. Innovation in machine learning and AI approaches at the n-of-1 level could help to accelerate the field.

**Author Summary:** The objective physiological and behavioral information from wearable and other smart devices is uniquely relevant to pregnancy. The objectives of this study were to: 1) describe the individual-level variability of pregnancy self-reported symptoms and objective wearable measures; 2) determine whether this variability can be explained by pregnancy clinical conditions; and 3) determine whether pregnancy self-reported symptoms are associated with objective wearable measures. Data are from the Better Understanding the Metamorphosis of Pregnancy study, which followed individuals from preconception to three-months postpartum. Participants (18-40 years) used an Oura smartring, a Garmin smartwatch, and a Bodyport Cardiac Scale alongside a study app to track self-reported symptoms. High heterogeneity was observed in self-reported pregnancy symptoms, and objective measures such as heart rate variability, activity and sleep over pregnancy that were dissimilar to the population average of these measures. Pregnancy clinical conditions did not explain well the observed high variability in objective wearable measures while self-reported symptoms were weakly correlated with objective wearable measures over pregnancy. In sum, high heterogeneity and complexities of associations between subjective experiences and objective measures from wearables across pregnant individuals pose challenges for researchers. Innovation in machine learning and AI individual level approaches will help to accelerate the field.

## Introduction

There is an unacceptable lack of evidence-based knowledge of and support for the pregnancy experience, internationally. Accordingly, there is unnecessary maternal morbidity and mortality associated with ineffective tools to identify early enough when a pregnant individual is at risk for a serious complication[1,2]. We have not until recently had the technology to adequately measure the needed scope and resolution of the dynamic pathophysiological processes that occur during pregnancy in near real time[3]. The paucity of existing research on pregnancy experiences, symptoms, related complications and conditions stems largely from un-representative study samples of pregnant individuals and uses aggregate approaches to characterize study data that often comprises subjective recollections of symptoms. The ‘aggregate approach’ combines individual level data and finds average trends within the group. This approach conveys how the average individual experiences pregnancy, in the context of the specific study sample used. There are a number of challenges with the aggregate approach that apply widely to the study of all diseases [4], but are uniquely relevant to pregnancy experiences that result from complex interactions between pre-pregnancy individual characteristics and proximal perinatal environments[5–8].

The expansion of the digital health technology (DHT) space over the past decade has given rise to novel, remote tools that can be worn on the person including wearables and smartphone apps, herein referred to as ‘personal DHTs’. Personal DHTs can be used to capture rich, semi-continuous multimodal (of different types) health-related information[9] that is uniquely relevant to pregnancy[10]. Consumer-grade wearables are capable of tracking semi-continuous measures of physiology (e.g. heart rate (HR), heart rate variability (HRV), body temperature, blood pressure, and oxygen saturation), behavior (e.g. activity, social activity, relative location, and sleep duration and quality) and metabolic information (e.g. weight, body fluid volumes) that include many in-clinic objective measures of health and additional measures not routinely captured that could provide objective measures of risky pregnancy-related symptoms[9,11]. Research approaches involving the collection of such data and in the use of machine learning and artificial intelligence (AI) techniques are showing promise for the personalized objective detection of symptoms such as gait[12], cognition[13], and mood[14] in other disease populations. The value in such data capture is in its objective nature, limiting reliance on subjective recollections of symptoms, ability to be captured outside healthcare settings, and in that it could encompass broad domains (pathophysiological, environmental, and psychosocial) that affect pregnancy experiences. Further, personal DHTs could enable the personalized monitoring (at the n-of-1 level) of pregnancy experiences and symptoms and be returned back to the user in near real time.

With the ubiquity of the smartphone[15], and the increasing popularity of wearable smart devices[16] such as the smartwatch and the smart ring, the use of such tools for monitoring pregnancy-related signs and symptoms and greatly transforming current knowledge of this important metamorphosis is a potential reality. While the body of research involving personal DHTs in pregnancy is small, a number of small preliminary studies have shown potential utility in using wearables for tracking activity, sleep, and physiologic metrics such as HR and HRV as predictors of pregnancy-related outcomes such as preterm birth and delivery readiness[17–23].

The aim of this paper is to elucidate the variability of the pregnancy experience at the n-of-1 level using multimodal personal DHT data from the Better Understanding the Metamorphosis of Pregnancy (BUMP) study - a US-based, participant centric digital health research study that follows individuals from preconception to the fourth trimester (three-months postpartum). Specifically, the study objectives were to: 1) characterize individual level pregnancy self-reported symptoms and objective wearable features compared to the aggregate; 2) determine whether pregnancy self-reported symptoms and objective wearable features can differentiate pregnancy-related conditions; and; and 3) describe associations between self-reported symptoms and objective wearable features.

## Results

The BUMP study enrolled 524 participants. This analysis included 406 participants who had at least one data point across the features examined and completed the study through birth. Characteristics of the sample can be found in Supplemental (Supp) Table 1. Participants were mostly white, 87% had a bachelor’s degree or higher, 82% were currently employed and working, and 34% were of advanced maternal age (35-40 years). In this study sample, there were 65 participants that experienced gestational hypertension, 61 that experienced postpartum depression, 43 that experienced preterm birth, 41 that experienced gestational diabetes, 39 that experienced postpartum hemorrhage, and 40 that experienced preeclampsia (3 of which experienced eclampsia). The distribution of body mass index across participants can be found in Supp. Figure 1.

### Objective 1 - Characterize individual level pregnancy self-reported symptoms and objective DHT features compared to the aggregate using data from personal DHTs

Figure 1 shows the proportion of participants reporting moderate to severe self-reported symptoms. In the aggregate, participants reported the following trends in self-reported pregnancy symptoms: increasing swelling, shortness of breath, gait changes and cognitive disturbances from the first to third trimesters, higher rates of fatigue during the first and third trimesters compared to the second trimester, higher rates of nausea and vomiting during the first trimester compared to all other trimesters, and increasing mood disturbances, peaking in the fourth trimester. Supp Figures 2a-g includes individual level raw data (coloured lines) and cohort averages (black line) across self-reported symptoms. On average, there were unique contours in self-reported symptoms over the four trimesters; however, there was high variability in self-reported symptoms at the n-of-1 level that show dynamic changes within individuals over pregnancy and into the postpartum period (Supp Figures 2a-g).

**Figure 1.**
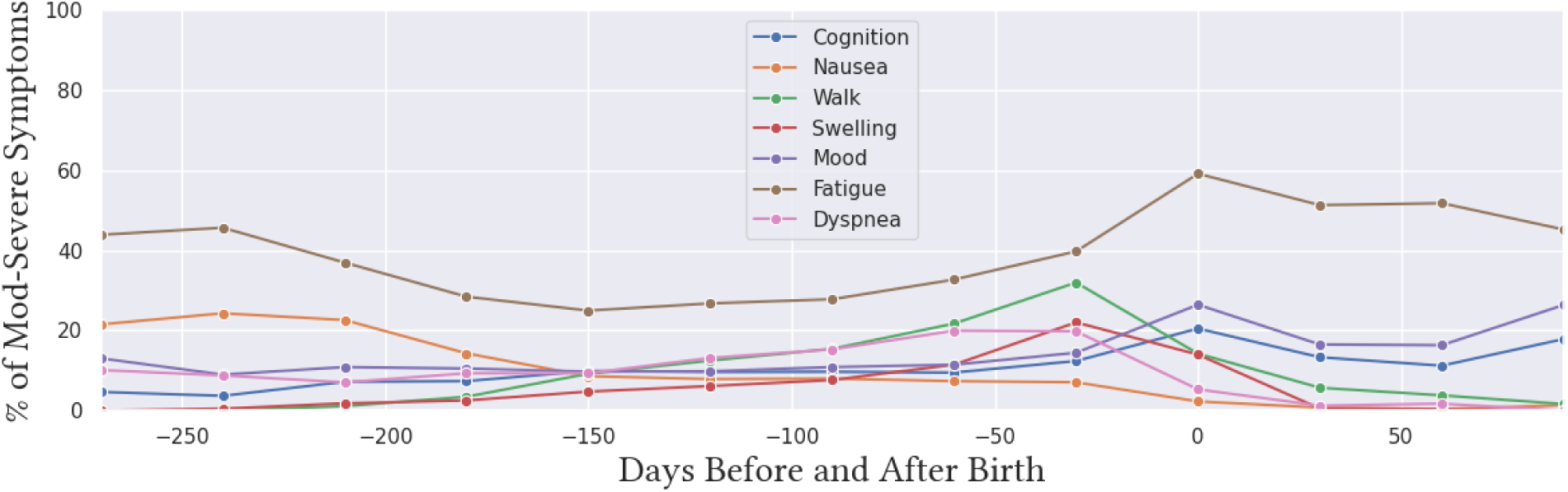
Proportion of participants reporting moderate to severe symptoms (nausea, shortness of breath, gait, swelling, fatigue, cognition and mood) over the four trimesters of pregnancy.

Similarly, cohort averages of objective features show unique contours over pregnancy (Figure 2-4). For example, on average, HR increases during the first two trimesters, and starts to decrease during the third trimester leading up to birth, while peripheral fluid volume steadily increases into the third trimester. HRV lowers with gestational age, but starts to increase leading up to delivery. The paradoxical inflection of HRV leading up to delivery is observed in the aggregate; however, is highly variable at the n-of-1 level, even among healthy pregnant individuals (participants who were not overweight, or obese, are under 35 years of age, and had no pre-existing medical conditions prior to pregnancy, or during pregnancy) (Figure 5). As observed in the self-reported symptom data, there is also high variability in objective feature data at the n-of-1 level both at baseline and over the course of pregnancy (Figures 2-4 and Supp Figure 3).

**Figure 2.**
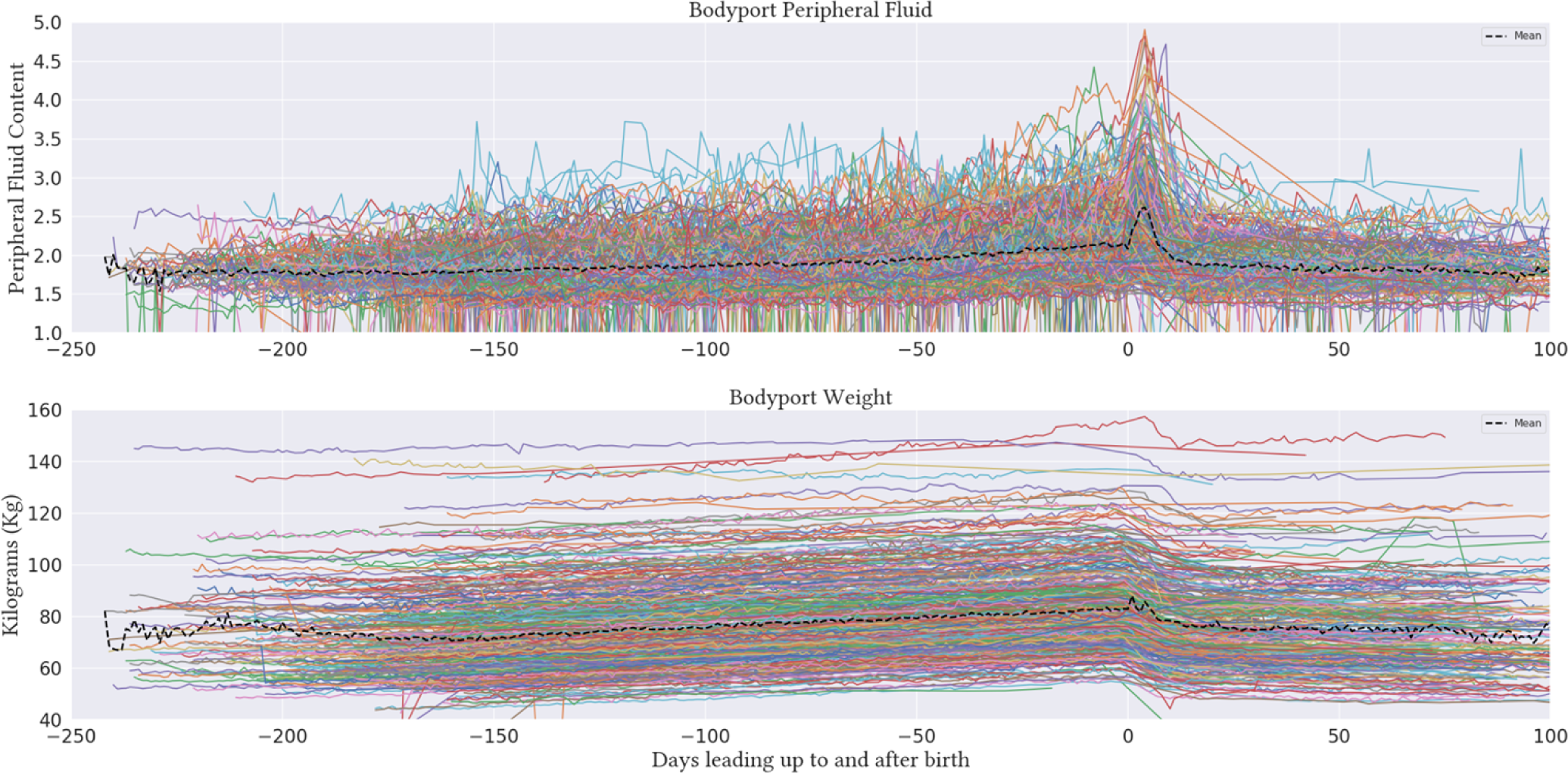
Individual level data (coloured lines) and cohort averages (black lines) for Bodyport peripheral fluid and weight over the four trimesters of pregnancy.

**Figure 3.**
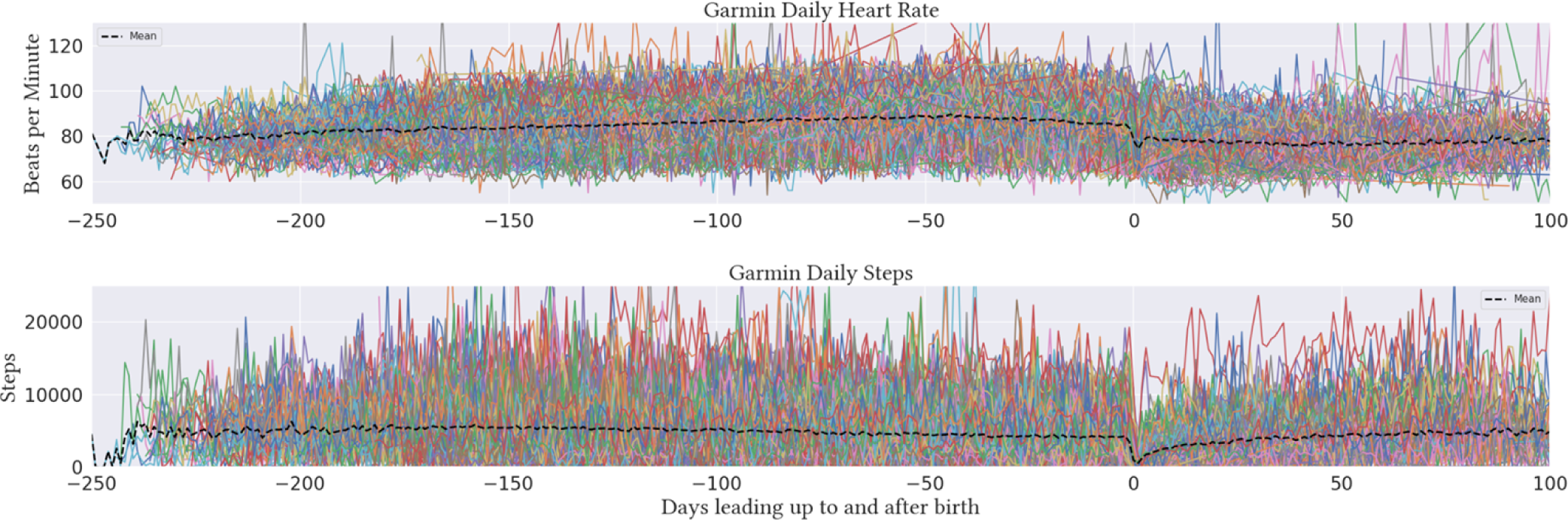
Individual level data (coloured lines) and cohort averages (black lines) for Garmin heart rate, respiratory rate, and activity, over the four trimesters of pregnancy.

**Figure 4.**
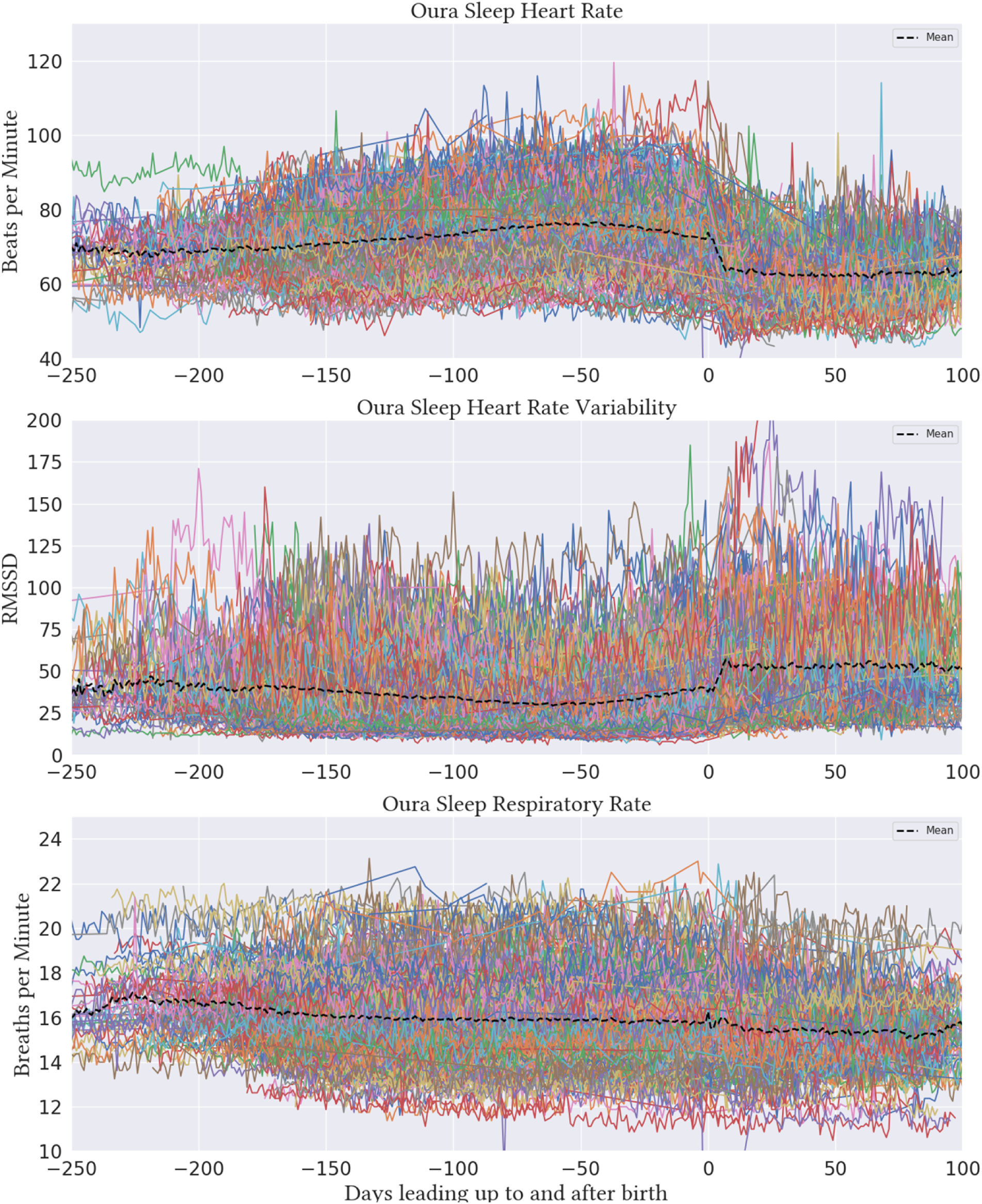
Individual level data (coloured lines) and cohort averages (black lines) for Oura night time heart rate, heart rate variability, and respiratory rate over the four trimesters of pregnancy.

**Figure 5.**
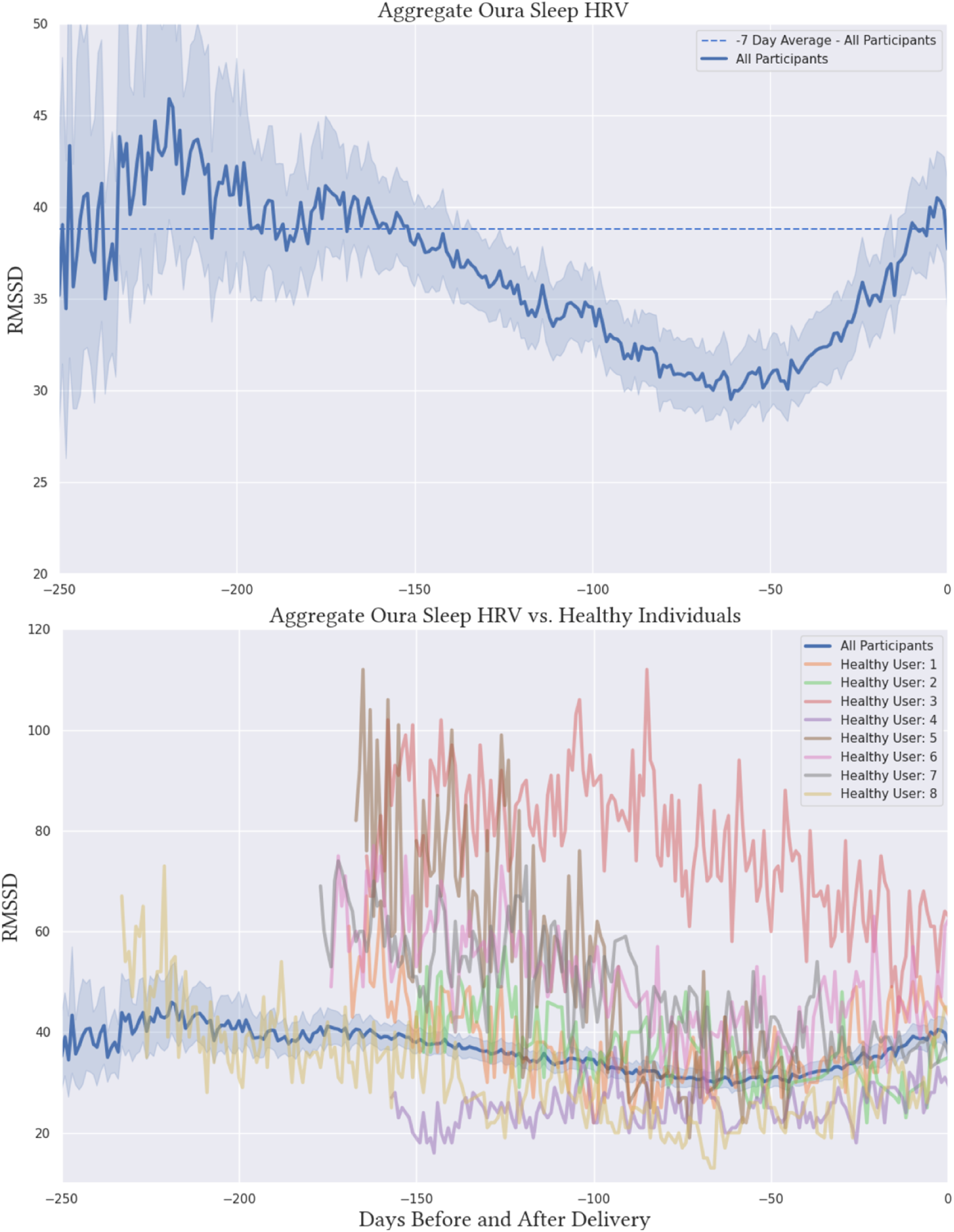
Average Oura nighttime HRV and n-of-1 level trends for eight unique healthy participants over pregnancy.

### Objective 2 - Determine whether pregnancy self-reported symptoms and objective features can differentiate pregnancy-related conditions

Supp Figures 4a-c include boxplots of each objective feature across different clinical condition groups, and the entire cohort. There was a high degree of overlap in the distribution of each objective feature across the groups, indicating that the observed heterogeneity was not explained well by clinical conditions.

Supp Figures 4-7 present cohort averages and 95% CIs of objective features and self-reported symptoms among healthy participants (n=26) and those that developed specific conditions over pregnancy. As expected, differentiation between the groups is observed for weight, particularly among the healthy group compared to the other condition groups as the healthy group was selected to only include normal weight individuals (Supp Figure 5).

Higher average night time HR and RR was observed in the clinical condition groups compared to the healthy group (Supp Figures 6a and b). Little differentiation between the groups was observed for average night time HRV and sleep efficiency (Supp Figures 6c and d). Cohort averages of daytime objective features measured by Garmin also showed little differentiation between the groups over pregnancy (Supp Figures 7a and b) with the exception of higher observed step count in the healthy group.

### Objective 3 - Describe associations between objective DHT features and subjective self-reported symptoms

Mean Spearman correlations across the examined self-reported symptom and objective feature pairs were close to 0 for all of the examined subjective/objective pairs with the exception of self-reported swelling and peripheral fluid measured from the Bodyport scale (Supp Table 2). Figure 6 shows individual level Spearman correlation strengths across all unique participants for two symptoms (swelling and shortness of breath). Self-reported swelling was correlated with peripheral fluid status in most participants (Figure 6). However, across all of the other self-reported symptoms - objective feature pairs, there was high variability in correlation strength across unique participants. In particular, there was a similar proportion of participants showing both positive and negative correlations between all of the examined feature pairs (Figure 6, Supp Figures 8a-f).

**Figure 6.**
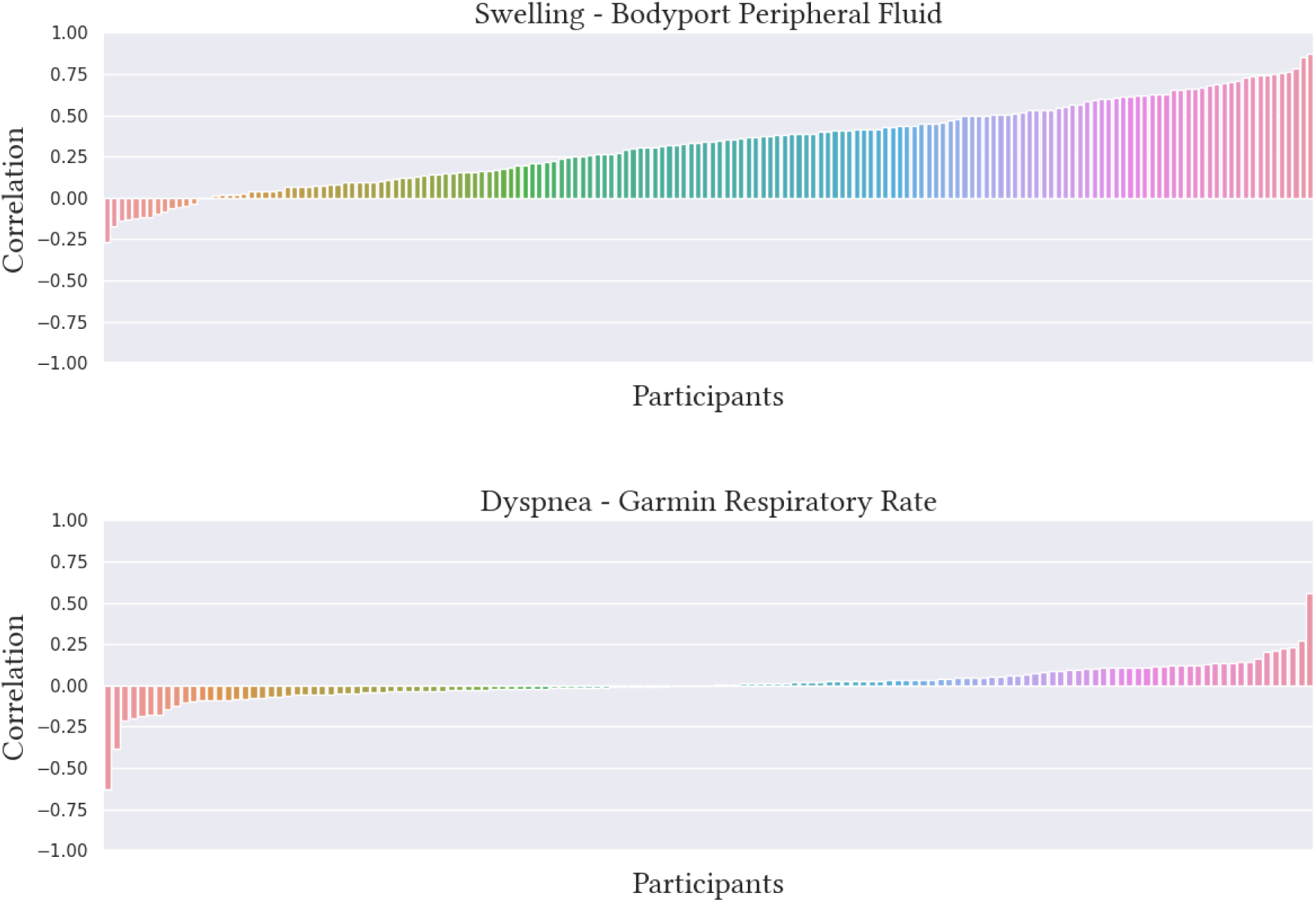
Individual level correlations between daily self-reported swelling and Bodyport Peripheral Fluid (top) and daily self-reported shortness of breath (dyspnea) and Garmin respiratory rate (bottom)

## Discussion

Self-reported symptoms and objective features reflective of physiologic, metabolic and behavioral changes over pregnancy, are markedly heterogeneous across individuals. While on average, pregnant individuals show unique contours of certain subjective self-reported symptoms and objective features from wearable devices, these patterns are misleading for many unique individuals. In particular, aggregate nighttime HR and RR were higher among individuals in pregnancy related-condition groups compared to healthy individuals - a difference that was observed early in pregnancy through birth, while HRV steadily decreased with gestational age, but showed an inflection at approximately nine weeks leading up to delivery. This phenomenon, that could reflect an individual’s physiological readiness for delivery, has been observed in the aggregate among other studies[18,23,26]. However, at the n-of-1 level, this aggregate trend is highly heterogeneous, even among healthy pregnant individuals.

Personal DHTs offer exciting opportunities to capture individual level high-resolution health-related information across multiple domains of health that could be used to glean a deeper insight into pregnancy pathophysiological changes and potential targets for early intervention. One of many values in leveraging the full richness of n-of-1 multimodal DHT data is in finding enough individuals who follow similar trajectories (e.g., the digital doppelganger) that might form new archetypes. These archetypes can then be used as the foundation for the development of early interventions. This is in contrast to the classical aggregate approach of finding groups of individuals who look similar to each other on average. These preliminary data demonstrate the potential challenge in identifying such archetypes owing to the observed high heterogeneity.

Subjective experiences of symptoms are limited by information biases. Objective measures of health enable an unbiased measure of an underlying biological or psychological process that could be used to identify when certain symptoms are becoming risky before the subjective experience arises. The degree of utility of objective measures of health to strengthen the subjective symptom relies on the nature of the symptom itself. For example, the subjective experience of swelling could be argued as an easier symptom to notice due to clear physical changes in the body as opposed to the subjective experience of cognition or mood that relies on individual level perceptions. These examples can be seen as differing examples of ground truths. Indeed, we demonstrated here that there was a more consistent concordance between self-reported swelling and the objective feature of body peripheral fluid volume among pregnant individuals compared to the other objective and subjective pairings. This finding may also highlight that certain objective features are more confounded than others. In line with this, across most of the selected self-reported subjective and objective features, there was high heterogeneity in correlation strength across participants with an almost equal number of participants showing positive and negative correlations. This finding highlights the challenges and complexity in leveraging single objective feature data to enhance subjective symptoms.

### Limitations

This paper was intended to highlight proof-of-concept, descriptive examples of raw, minimally processed DHT data to convey the complexity of this information in describing the pregnancy symptom experience. This complexity will create challenges at the quantitative level, while some personalized methodologies might aid in uncovering trends, such as subgroup clustering in the search for digital doppelgangers. However, this was out of scope for this paper. In this study, we did not have access to raw photoplethysmography (PPG) data; therefore, could not explore morphological features of the PPG waveform that has been shown to potentially be more useful for monitoring maternal health states compared to HRV[22]. The self-reported symptom and objective feature pairings selected for this preliminary analysis may not be the right pairings or may be too difficult to quantify consistent associations due to confounding factors. The healthy comparison group was selected to only include healthy weight participants, >35 years of age, and with no prior or current medical conditions. The fact that this group did not include overweight or obese participants may in part explain the higher aggregate trend in nighttime HR and RR in the condition groups compared to this comparison group. The BUMP study sample is non-generalizable to other populations, and in particular to underrepresented populations as the proportion of participants from non-white ethnicities and races was low.

### Conclusions

Personal DHTs offer a novel lens through which to examine the pregnancy experience at high resolutions, across broad multimodal domains and in objective ways. However, high within-person heterogeneity and the plethora of confounders of objective wearable signals highlight the danger in reliance on the aggregate approach to informing individual level pregnancy experiences. Further, the use of objective wearable signals to strengthen the subjective self-reported symptom are also likely impeded by within person confounding. These complexities pose challenges for researchers and underscore the importance of individualized approaches to data capture and analysis.

### Future Directions

Wearable data is noisy. Large sample sizes will be required to accommodate high individual level heterogeneity. Innovation in machine learning and AI approaches that are able to embrace the chaotic nature of these data will be needed to accelerate the field. As opposed to using analytic approaches that attempt to control for noise, alternative methods that aim to find signals within the individual level heterogeneity may be more effective. In the context of the pregnancy experience, exploration into individual level rhythms of multimodal signals that can be expected during pregnancy, and for a unique individual, and then deviation from these rhythms could reflect new ways of conceptualizing personalized risk states.

## Materials and Methods

### Design Overview

Data were derived from the BUMP study[3]. Please refer to the BUMP published protocol for study design details[3]. Briefly, the BUMP study is a longitudinal participant-centric digital health observational study that followed individuals from preconception to three-months postpartum. Participants were recruited from a variety of channels including patient provider portals (Sema4), social media advertisements, and community health clinics. Individuals between 18-40 years of age and who were actively trying to get pregnant were invited to participate in a conception cohort (BUMP-C). Individuals 18-40 years of age and up to 15 weeks pregnant with no intention to terminate their pregnancy were invited into the main BUMP cohort and followed until three-months postpartum. Participants in the BUMP-C cohort were followed for six months, and if they became pregnant during this time, were invited to join the BUMP cohort. This analysis only includes data from the BUMP cohort. Participants in the BUMP cohort were provisioned with an Oura smart ring, a Garmin smartwatch, and a Bodyport Cardiac Scale and were asked to use these devices continuously throughout their participation alongside a BUMP study app. The BUMP app prompted participants with daily and intermittent symptom surveys and active tasks. Participants’ electronic pregnancy-related electronic health record (EHR) data over pregnancy was also collected[3]. The BUMP study was ethically approved by the Institutional Review Board, Advarra (Pro00047893). E-consent was obtained from all participants in the study app.

### Measures

The BUMP study collected many different types of multimodal wearable and smartphone data described in detail elsewhere[3]. For this specific analysis, we examined data from select objective features from the study wearable devices and distinct self-reported symptoms from app surveys as outlined below.

#### Wearable and smart devices

The Oura Ring 2 (https://ouraring.com/) is a lightweight, durable titanium device worn on the finger that includes a temperature sensor, a gyroscope, a 3D accelerometer and an infrared optical pulse sensor. The device collects a variety of physiological, activity and sleep features. For this analysis the following features were examined: average night time HR, average night time HRV calculated from the rMSSD method, average night time respiration rate (RR), night time body temperature deviation (average temperature relative to baseline normal range), and sleep efficiency (proportion of sleep period spent asleep).

The Garmin Venu Sq Smartwatch ((https://www.garmin.com/) includes a 3-axis accelerometer, 3-axis gyroscope, Optical HR monitor, Altimeter and a Vibration motor. The device is made of a lightweight material similar to that used in many sports watches and is able to measure physiologic and activity features. For this analysis, the following features were examined: average daily HR and steps.

The Bodyport Cardiac Scale (https://bodyport.com/) comprises a physical platform on which the user stands with bare feet. Four electrodes located on the top surface of the platform are used to obtain three biological signals from the user’s body. The first is a passive electrical signal and is similar in origin to an electrocardiograph. The second reflects pulsatile blood flow and is determined by measuring small changes in the electrical resistance through the legs. The third signal measures blood flow velocity through the Aorta and reflects mechanical function of the heart. From these signals, the scale measures several cardiovascular and metabolic markers. For this analysis, we examined weight and body peripheral fluid levels.

#### App Surveys

Participants were prompted daily to complete a nine-item symptom survey about pregnancy-related experiences that asked: “In the past day, have you noticed any symptoms? (e.g., nausea/vomiting, fatigue, swelling, memory, mood, shortness of breath, the way you walk)? (Yes/No). If participants indicated no to the initial screening question, the survey ended. If participants responded yes to the original screening question they were provided with each symptom and asked to rate its severity on a likert scale from 1=none to 7=severe. This survey was designed by 4YouandMe with the intention of following key symptoms that have the potential to be followed by objective wearable and smartphone data. Participants also completed a one time in-app socio-demographic and medical history survey and a post-birth survey over the phone with research staff that captured self-reported information on diagnosis of pregnancy-related conditions[3].

Pregnancy related conditions were measured using a multi-source data approach. Participants completed a post-birth survey between one to three months postpartum over the phone with a research coordinator that asked participants to self-report on pregnancy-related conditions and complications including gestational hypertension and diabetes, preeclampsia, eclampsia, toxemia, preterm birth, postpartum depression and others. This information was enriched using participant EHR data that was captured from Sema4’s patient platform that requests users’ digital consent to access their EHR data[3].

#### Other App Active Tasks

Participants completed in-app intermittent active tasks. For this analysis, the Cognition Kit N-Back Task (N-Back)[24] - a task of attention and working memory that prompts participants to respond when a presented symbol is the same as the symbol presented from two previous trials (prompted within the app every three days) and the CANTAB Emotional Bias Task (EBT)[25] - a task of perceptual bias of facial emotions that prompts the participants to indicate whether human faces appear happy or sad (prompted in the app every week) were used.

### Analysis

Participants were included in this analysis if they had given birth and had at least one data point across the different features used including: Oura Sleep data, Bodyport non-cardiac and balance data, Garmin Daily data, a baseline socio-demographic survey, medical history survey and a daily symptom survey. Data are largely presented in individual level raw format with minimal processing for the purposes of describing their complexity. Cohort averages were estimated by calculating the daily average of each variable before and after delivery. Participants who had fewer than ten data points in each variable were excluded from aggregate analyses.

Individual level differences from the cohort average can be expected from many confounding factors such as pre-existing or newly developed conditions in pregnancy or other individual characteristics such as age and weight. Therefore, cohort averages were stratified by pregnancy clinical conditions including participants with pre-eclampsia, gestational hypertension, gestational diabetes, postpartum hemorrhage, postpartum depression, and a selected healthy group to include participants who were >35 years of age, whose prepregnancy weight was not overweight or obese (body mass index <25) and had no known pre-existing clinical conditions, or any clinical conditions that newly developed over pregnancy.

Pairs of associations between each of the daily self-reported symptoms and selected objective features, and active tasks that could be associated (e.g., self-reported shortness of breath and respiratory rate, or self-reported mood and emotional bias scores) were examined. Individual level associations were estimated using Spearman Correlations. Pairing events were identified in each unique participant’s time series. For semi-continuous and daily data (Garmin, Oura, Bodyport and daily symptom survey data) pairing events were defined as days where a self-reported symptom and objective feature data event were present. In-app active tasks were captured every few days to weekly; therefore, pairing events for these instances were defined as periods where self-reported symptom data was present in the week preceding an active task data event. The self-reported symptom responses were averaged over the week preceding the active task data event. If there was less than a week’s worth of self-reported symptom data in the week preceding the active tasks data event, missing days were omitted from the mean calculation. Participants were included in correlation analyses if they had at least ten pairing events. Data were analyzed using python 3.6.10 with pandas 0.25.3, NumPy 1.19.5, SciPy 1.4.1, scikit-learn 0.22.2, matplotlib 2.2.5, and Seaborn 0.11.2 modules.

## Data Availability

Among participants who opt-in, coded study data from the BUMP study participants is available on the Synapse platform (synapse.org) at Sage Bionetworks (https://sagebionetworks.org) and can be freely accessed by any researcher who becomes ‘qualified’ by becoming a registered and certified Synapse user (https://help.synapse.org/docs/User-Account-Tiers.2007072795.html), and by meeting the specific conditions of use that require submitting an intended data use statement alongside an IRB approved protocol. The BUMP specific Synapse Project page can be found here: https://www.synapse.org/#!Synapse:syn25953345/wiki/616547 among registered Synapse users.

## Acknowledgements

We would like to extend sincere gratitude to the current and future BUMP and BUMP-C participants for your dedication to this work to help other women transitioning to and experiencing pregnancy.

## Supporting Information

**Supp. Table 1. Study sample characteristics**

**Supp. Table 2. Mean Spearman correlation across participants for select subjective self-reported symptoms and objective features pairs**

**Supp. Figure 1. Body Mass Index (BMI) distribution across participants**

**Supp. Figure 2a. Individual level (coloured lines) and cohort averages (black lines) of subjective self-reported swelling over pregnancy**

**Supp. Figure 2b. Individual level (coloured lines) and cohort averages (black lines) of subjective self-reported shortness of breath over pregnancy**

**Supp. Figure 2c. Individual level (coloured lines) and cohort averages (black lines) of subjective self-reported gait changes over pregnancy**

**Supp. Figure 2d. Individual level (coloured lines) and cohort averages (black lines) of subjective self-reported low mood over pregnancy**

**Supp. Figure 2e. Individual level (coloured lines) and cohort averages (black lines) of subjective self-reported nausea over pregnancy**

**Supp. Figure 2f. Individual level (coloured lines) and cohort averages (black lines) of subjective self-reported fatigue over pregnancy**

**Supp. Figure 2g. Individual level (coloured lines) and cohort averages (black lines) of subjective self-reported changes in cognitive function over pregnancy**

**Supp. Figure 3 - Individual level data (coloured lines) and cohort averages (black lines) for Oura ring sleep efficiency and temperature deviation over the four trimesters of pregnancy**

**Supp Figure 4a - Box plots for weight and peripheral fluid measured from the Bodyport Cardiac Scale across clinical groups, healthy participants and all enrolled participants in the BUMP cohort**

**Supp Figure 4b - Box plots for heart rate and steps measured from the Garmin smartwatch across clinical groups, healthy participants and all enrolled participants in the BUMP cohort**

**Supp Figure 4c - Box plots for heart rate, heart rate variability, respiratory rate, sleep efficiency and body temperature deviation measured from the Oura smartring across clinical groups, healthy participants and all enrolled participants in the BUMP cohort**

**Supp Figure 5 - Cohort averages and 95% Confidence Intervals (CIs) for weight and peripheral fluid content measured from the Bodyport Cardiac Scale across pregnancy condition and healthy groups over the four trimesters**

**Supp Figure 6a - Cohort averages and 95% Confidence Intervals (CIs) for night time heart rate measured from the Oura ring across pregnancy condition and healthy groups over the four trimesters**

**Supp Figure 6b - Cohort averages and 95% Confidence Intervals (CIs) for night time respiratory rate measured from the Oura ring across pregnancy condition and healthy groups over the four trimesters**

**Supp Figure 6c -Cohort averages and 95% Confidence Intervals (CIs) for night time heart rate variability measured from the Oura ring across pregnancy condition and healthy groups over the four trimesters**

**Supp Figure 6d - Cohort averages and 95% Confidence Intervals (CIs) for sleep efficiency (proportion of time spent asleep) measured from the Oura ring across pregnancy condition and healthy groups over the four trimesters**

**Supp Figure 7a - Cohort averages and 95% Confidence Intervals (CIs) for heart rate measured from the Garmin watch across pregnancy condition and healthy groups over the four trimesters**

**Supp Figure 7b - Cohort averages and 95% Confidence Intervals (CIs) for daily steps measured from the Garmin watch across pregnancy condition and healthy groups over the four trimesters**

**Figure 8a. Individual level correlations between daily subjective cognitive function and Nback score**

**Figure 8b. Individual level correlations between daily subjective gait and Bodyport Sway Velocity**

**Figure 8c. Individual level correlations between daily subjective mood and Active Task Emotional bias point**

**Figure 8e. Individual level correlations between daily subjective mood and Garmin Active Time**

**Figure 8f. Individual level correlations between daily subjective nausea and vomiting and Bodyport peripheral fluid volume**

